# Three Sibling Genes Involved in Genetic Risk for Lateral Epicondylopathy

**DOI:** 10.64898/2026.02.16.26346404

**Authors:** Kaitlyn S. Burns, Stuart K. Kim, William Denq

**Affiliations:** College of Medicine-Tucson, University of Arizona, Tucson, AZ; Dept. Dev. Bio., Stanford University Medical Center, Stanford, CA; Department of Emergency Medicine, University of Arizona, Tucson, AZ

## Abstract

**Objectives:** To screen the entire genome for genes associated with risk for lateral epicondylopathy and improve understanding of underlying biological mechanisms and inform future research aimed at risk stratification and personalized prevention and treatment strategies.

**Methods:** A genome-wide association study was conducted using UK Biobank data. Lateral epicondylopathy cases were identified based on electronic health records from individuals of European ancestry. Logistic regression tested associations between single-nucleotide polymorphisms and disease status, adjusting for sex, age, height, weight and ancestry principal components. Previously-identified candidate genes from the literature were also tested for association with lateral epicondylopathy.

**Results:** Among 20,390 cases of lateral epicondylopathy, two loci reached genome-wide significance: one comprising 144 linked SNPs and one single SNP. The first locus, led by rs13127477 (p=7.7×10^−12^; OR 0.93, 95% CI 0.91 to 0.95), is located near three SIBLING genes (IBSP, MEPE and SPP1) involved in extracellular matrix remodelling at fibrocartilaginous entheses. The risk allele was associated with increased SIBLING gene expression, suggesting that excessive entheseal matrix remodelling contributes to disease susceptibility. The second locus was defined by rs138254824 (p=3.69×10^−8^; OR 3.42, 95% CI 2.23 to 5.25) near NEDD9 and TMEM170B. Previously reported collagen gene associations were not replicated.

**Conclusion:** In the first genome-wide screen for lateral epicondylopathy, two loci were identified. These loci provide insight regarding the pathophysiology of lateral epicondylopathy and a roadmap for preventing and treating this injury with personalized medicine.

**Summary Box:** *What is already known on this topic:* Lateral epicondylopathy is a common and disabling overuse tendon condition, yet its genetic basis has remained poorly characterised, with prior studies limited to small candidate gene analyses.

*What this study adds:* This study provides the first genome-wide association analysis of lateral epicondylopathy, identifying two risk loci on chromosomes 4 and 6 and implicating SIBLING genes (IBSP, MEPE, and SPP1) involved in entheseal extracellular matrix remodelling.

*How this study might affect research, practice or policy:* These findings offer new biological insight into disease susceptibility and challenge previously reported collagen gene associations.

## Introduction

Lateral epicondylopathy is an overload injury of the common extensor tendon at the lateral humeral epicondyle.^1^ It affects 1-3% of the general adult population and more than 50% of athletes performing repetitive or overhead upper-limb activities.^1^ It presents with insidious lateral elbow pain, tenderness over the lateral epicondyle, and pain with resisted wrist extension. The condition reflects a degenerative process termed angiofibroblastic tendinosis, characterised by fibroblast infiltration, vascular hyperplasia, disorganised collagen, and increased inflammatory cytokines.^2,3^

Despite its prevalence and functional impact, genetic susceptibility to lateral epicondylopathy remains poorly characterised. Previous studies have reported that candidate collagen genes (*col5A1* and *col11A1*) were risk factors associated with lateral epicondylopathy.^4,5^

Genome-wide association (GWA) analyses enable unbiased identification of genetic risk loci by testing millions of variants across the genome. Identifying genetic contributors to lateral epicondylopathy may improve understanding of disease pathophysiology and individual risk profiles, informing prevention and treatment strategies. Here, we report the first genome-wide association study of lateral epicondylopathy.

## Methods

### Study Population

Genotype data were obtained from the v3 release of UK Biobank.^6^ The UK Biobank electronic health care records were available for 471,929 individuals. Genotype data were imputed centrally by UK Biobank with IMPUTE2 using the Haplotype Reference Consortium and the UK10k + 1000GP3 reference panels.^7^ Metrics for quality control were established and then used to filter DNA variants by UK Biobank.^6^ Imputed SNPs were excluded if they had an IMPUTE2 info score <0.4.

### Phenotype Definition

Cases of lateral epicondylopathy were identified using International Classification of Diseases (ICD9 and ICD10) codes from hospital episode statistics and Read2/Read3 codes extracted from general practitioner clinical records (Table 1). Control participants were defined as all remaining UK Biobank participants without these diagnostic codes. Participants with missing covariates or poor-quality genotyping data were excluded prior to association testing. A total of 20,390 cases and 451,539 controls were included in the final analysis.

**Table 1:** Diagnostic codes for lateral epicondylopathy.

|  |  |  |  |
| --- | --- | --- | --- |
| Table 1: Diagnostic codes for lateral epicondylopathy |  |  |  |
|  | Code | Description | Number |
| ICD10 | M77.10 | Lateral epicondylitis, unspecified elbow | 3 |
| ICD10 | M77.11 | Lateral epicondylitis, right elbow | 3 |
| ICD10 | M77.12 | Lateral epicondylitis, left elbow | 575 |
| Read2 | N2132 | Lateral epicondylitis of the elbow | 8,213 |
| Read3 | N2132 | Lateral epicondylitis of the elbow | 22,654 |
|  |  | Total Unique Patients | 20,390 |

### Database quality control

Individuals were excluded if they were outliers based on genotyping missingness rate or heterogeneity, whose sex inferred from the genotypes did not match their self-reported sex, who withdrew from participation, or who were not of European ancestry. Restricting the analysis to individuals of European ancestry reduces confounding from population stratification, which occurs when systematic differences in allele frequencies between ancestral groups produce spurious associations. Overall, these filters resulted in excluding 7.9% of individuals (mostly due to the ancestry filter). Genetic variants that failed quality control procedures in any of the genotyping batches, that showed a departure from Hardy–Weinberg equilibrium of P < 10^−50^, that had a minor allele frequency < 0.001, or missing genotype rate > 5% were excluded. The determination of genetic ancestry was performed by principal component analysis computed centrally by UK Biobank, as previously described.^6^

### Genetic Association Analysis

The genome-wide association study (GWAS) was conducted using PLINK v2.0a.^8^ SNP associations were tested for association with lateral epicondylopathy with a logistic regression model using allele counts for typed and imputed SNPs. The model was adjusted for genetic sex, age, height, weight and race/ethnicity (using 15 principal components).The final number of SNPs that was analyzed was 17,541,396. To account for inflation due to population stratification, the genomic control parameter (λgc) was calculated (λgc = 1.057) and used to adjust P values accordingly. Genome-wide significance was defined as p < 5×10^−8^.

QQ and Manhattan plots were created using the qqman function in R. Regional association plots were generated for each locus with LocusZoom (http://locuszoom.org/; accessed December 1, 2025).^9^ Whether each SNP is an expression quantitative trait locus (eQTL) was queried using the Genotype-Tissue Expression (GTEx) Portal (https://gtexportal.org/home/). The genomic context of each SNP was investigated using RegulomeDB

(https://regulomedb.org/)^10^ Web tools. Summary statistics for all SNPs from the GWAS will be available at the NHGRI GWAS catalog (https://www.ebi.ac.uk/gwas/) upon acceptance of this manuscript.

## Results

### Identification of DNA variants associated with lateral epicondylopathy

We performed GWA analysis for lateral epicondylopathy with the UK Biobank cohort (471,929 individuals) using sex, weight, height, age and 15 principal components as adjustments. There were 20,390 cases and 451,539 controls (Table 2). The lateral epicondylopathy cohort was modestly older and had a slightly higher proportion of females than the control cohort.

**Table 2:**
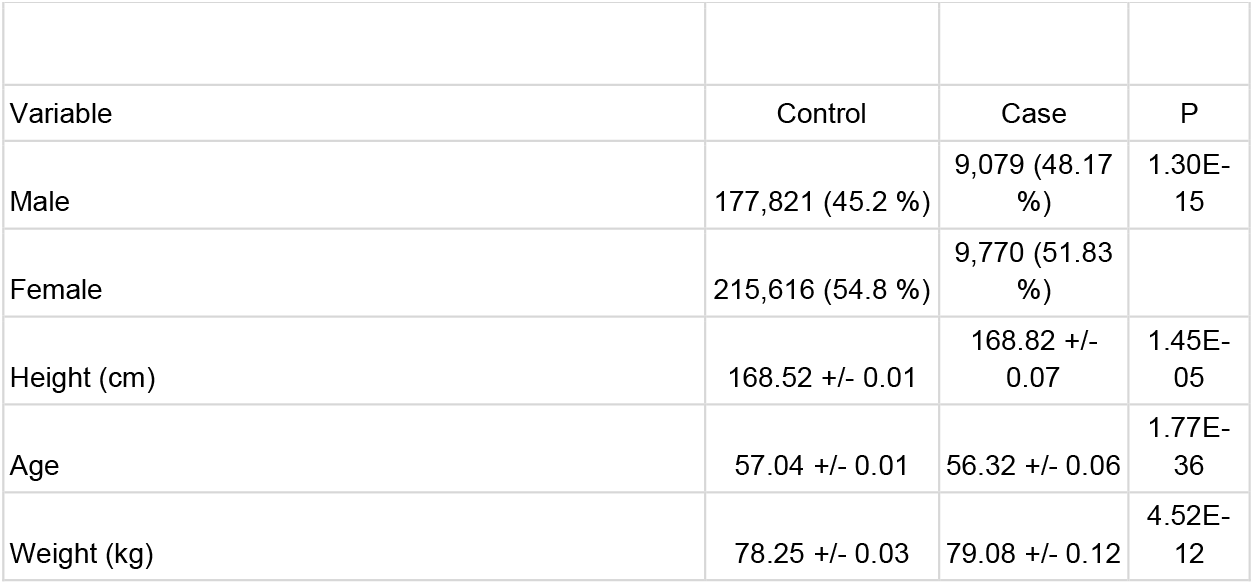
Demographic data for the UK Biobank cohort.

|  |  |  |  |
| --- | --- | --- | --- |
| Table 2: Demographic data for the UK Biobank cohort |  |  |  |
| Variable | Control | Case | P |
| Male | 177,821 (45.2 %) | 9,079 (48.17 %) | 1.30E-15 |
| Female | 215,616 (54.8 %) | 9,770 (51.83 %) |  |
| Height (cm) | 168.52 +/- 0.01 | 168.82 +/- 0.07 | 1.45E-05 |
| Age | 57.04 +/- 0.01 | 56.32 +/- 0.06 | 1.77E-36 |
| Weight (kg) | 78.25 +/- 0.03 | 79.08 +/- 0.12 | 4.52E-12 |

We compared the observed p-values to the distribution of p-values expected by chance in a Q-Q plot (Fig. 1). The black dots deviate from the red line for the lowest observed p-values in the upper right-hand corner, indicating that the observed association signals are significantly different from the signals that would be expected by chance.

**Fig. 1.**
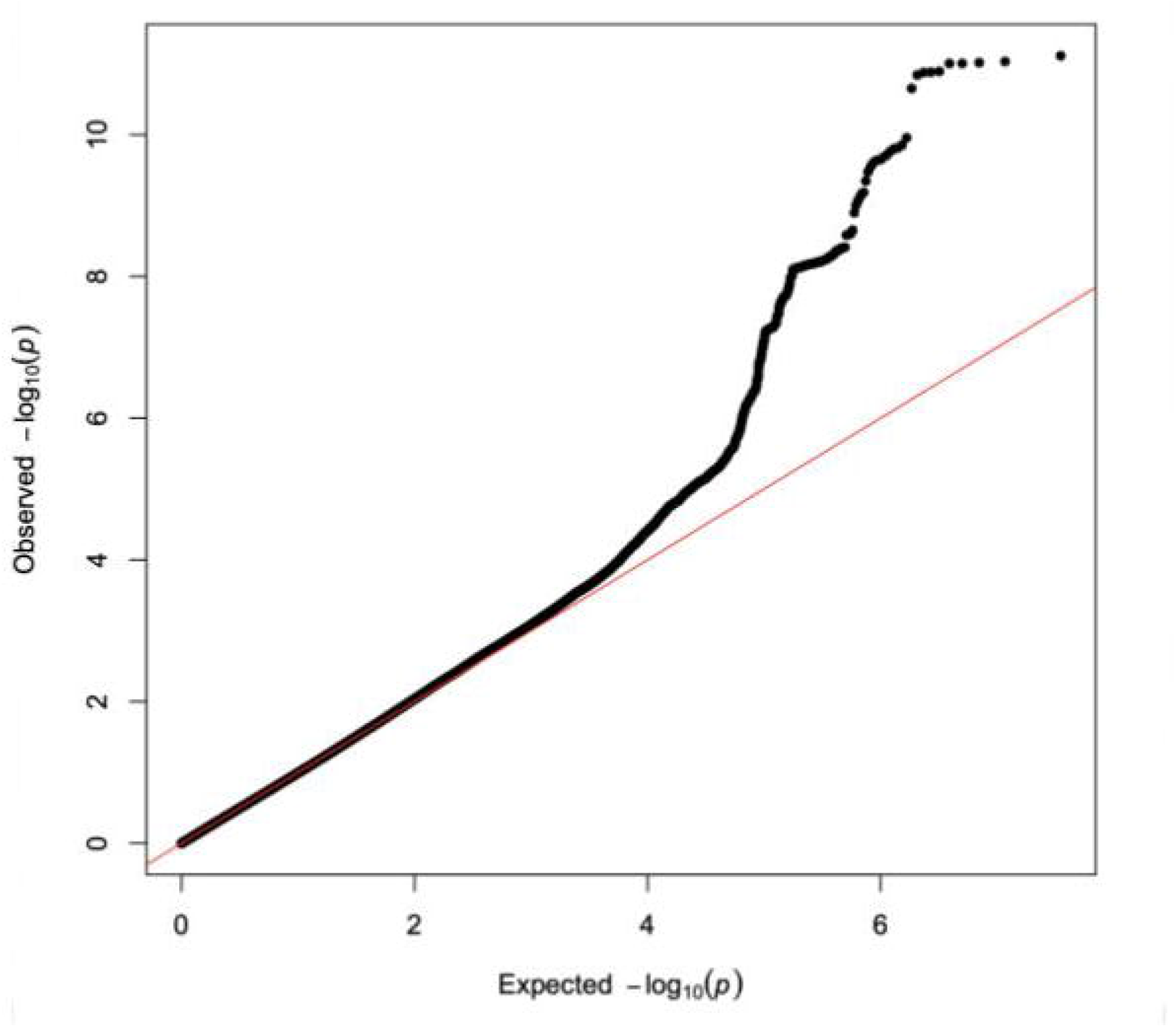
QQ plot with the observed p-values compared to those that would be expected by chance. The observed p-values and the p-values expected by chance (red line) are plotted as black and red dots, respectively. The observed p-values show a strong deviation from those that would be expected by chance.

The p-value for every SNP from the meta-analysis is shown in a Manhattan plot (Fig. 2). Using p = 5×10^−8^ as a cut-off for genome-wide significance, there were a total of 145 SNPs located in two loci on chromosomes 4 and 6 (Table 4, Sup. Table 1).

**Fig 2.**
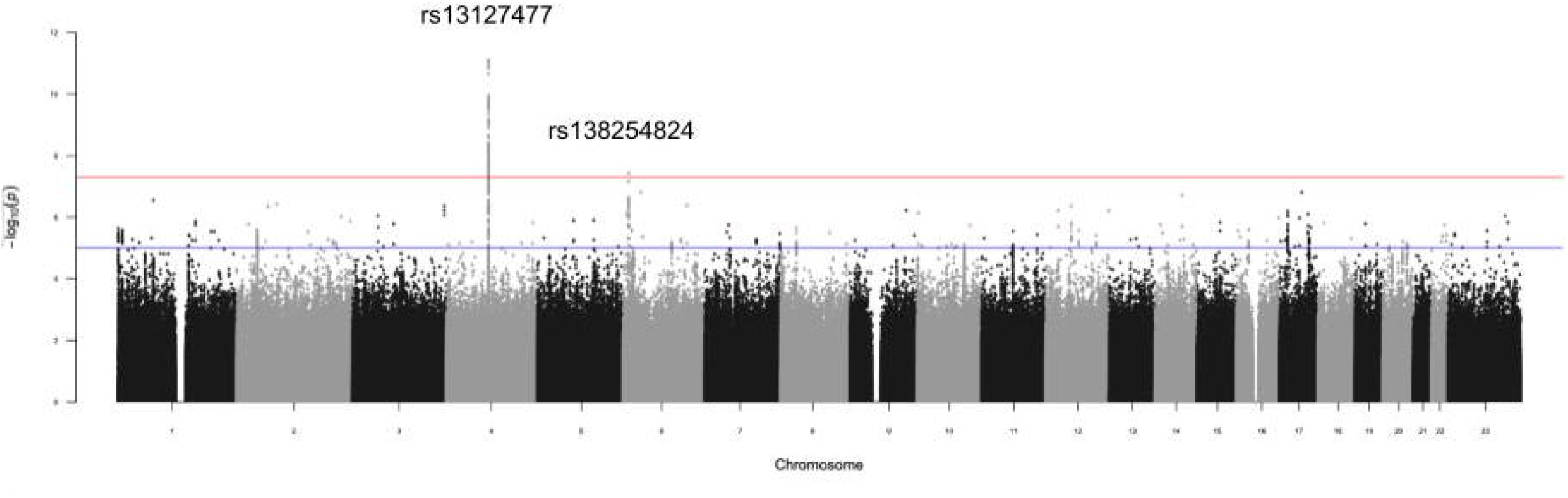
Manhattan plot for genome-wide association analyses of lateral epicondylopathy in the EUR ancestry group. The −log10 p-values for association with lateral epicondylopathy are plotted by genomic position with chromosome number listed across the bottom. The y-axis shows the −log10 p-value for their association with lateral epicondylopathy. The blue line represents suggestive genome-wide significance (p<1×10−5) and the red line represents genome-wide significance (p<5×10^−8^).

The locus on chromosome 4 consists of 144 SNPs that are in close proximity to each other. Of these, rs13127477 is the lead SNP as it shows the strongest association with lateral epicondylopathy, with a p value = 7.7×10^−12^. We tested whether the set of SNPs from this locus represents just one association, in which case one SNP could be causally associated with lateral epicondylopathy and the remainder might show an association because they are correlated (i.e. in linkage disequilibrium) with the causal SNP. The lead SNP rs13127477 might be a causal SNP or it may also be a tag for a causal SNP that is not observed. An alternative is that there may be more than one signal showing independent associations with lateral epicondylopathy. To distinguish between these possibilities, we determined whether any of the 143 SNPs retained an association with lateral epicondylopathy when conditioned on the genotype of rs13127477. We found that none showed an independent association. Thus, the

SNPs on chromosome 4 represent one linkage disequilibrium block with rs13127477 as the lead SNP. The A allele of rs13127477 has an odds ratio of 0.926 (0.906 to 0.946; 95% CI) for risk of lateral epicondylopathy compared to the G allele (Table 3). The SNP on chromosome 6 (rs138254824) showed a p value of 3.7×10^−8^. The T allele has an odds ratio of 3.4 (2.2 to 5.2; 95% CI) compared to the C allele, meaning that the T allele is associated with a higher prevalence of injury.

**Table 3:** Lead SNPS from the genome-wide screen for lateral epicondylopathy (p < 5E-8).

|  |  |  |  |  |  |  |
| --- | --- | --- | --- | --- | --- | --- |
| SNP | Genes | Chromosome | Position | EA | OR | p-value |
| rs13127477 | IBSP / MEPE / SPP1 | 4 | 88,806,268 | A | 0.926<br>(0.906 to 0.946) | 7.71E-12 |
| rs138254824 | NEDD9 / TMEM170B | 6 | 11,452,224 | T | 3.4294<br>(2.238 to 5.254) | 3.69E-08 |

Figure 3A shows a zoom plot for the association of SNPS on chromosome 4 with lateral epicondylopathy. The SNPs are located in the intergenic region between the *IBSP* and *MEPE* genes on the left, and the *SPP1* gene on the right. *IBSP* encodes integrin-binding sialoprotein that is produced by bone cells as a major structural protein of the bone extracellular matrix that mediates cell attachment.^11^ The *MEPE* gene encodes matrix extracellular phosphoglycoprotein, a protein involved in bone mineralization and phosphate homeostasis.^12^ *SPP1* encodes osteopontin, which is a key non-collagenous bone matrix protein that mediates attachment of osteoclasts to bone matrix.^13^ IBSP, MEPE and SPP1 are SIBLING genes (Small Integrin-Binding Ligand N-linked Glycoprotein) involved in extracellular matrix remodeling at the fibrocartilaginous enthesis.^14^

**Fig 3.**
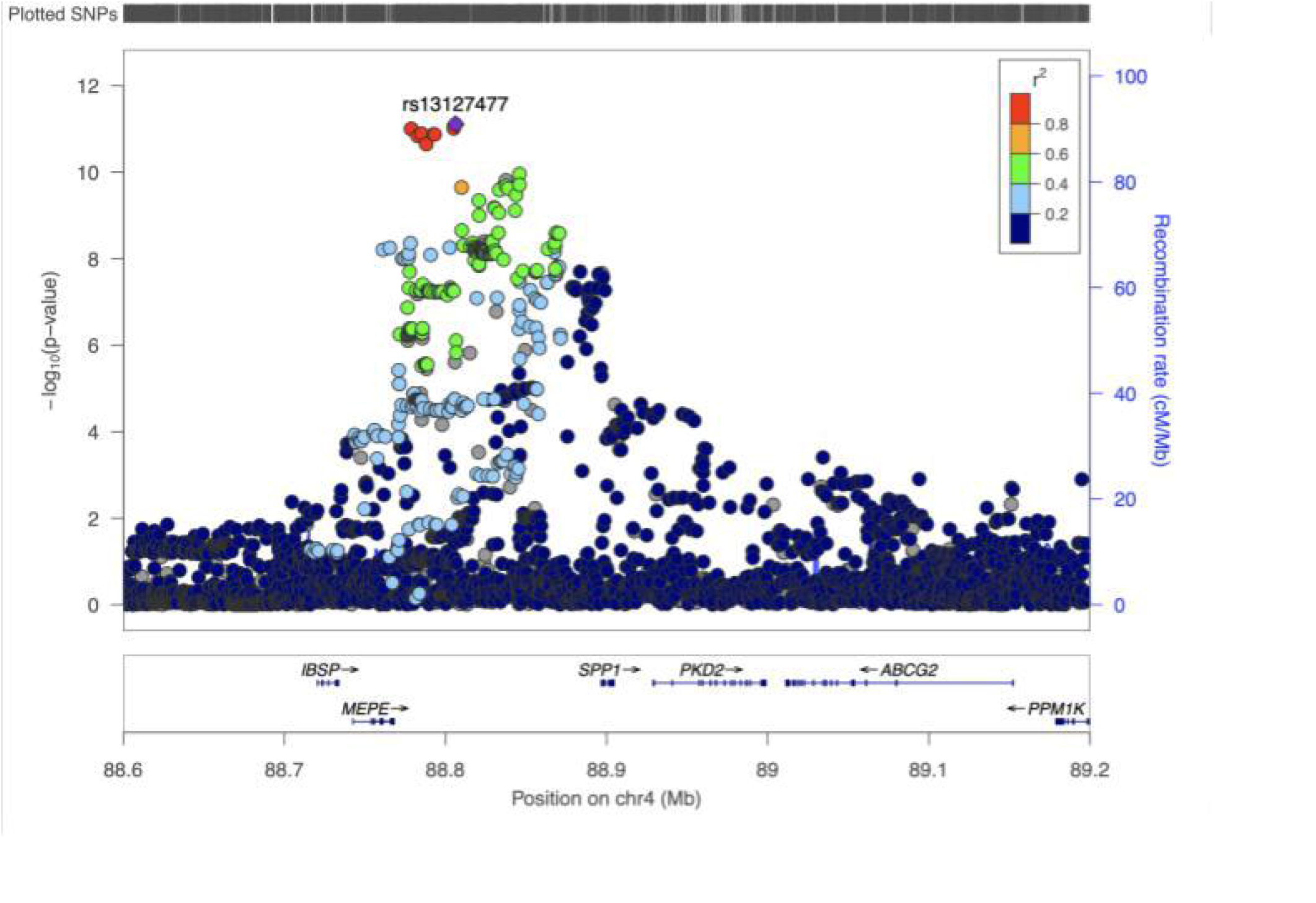
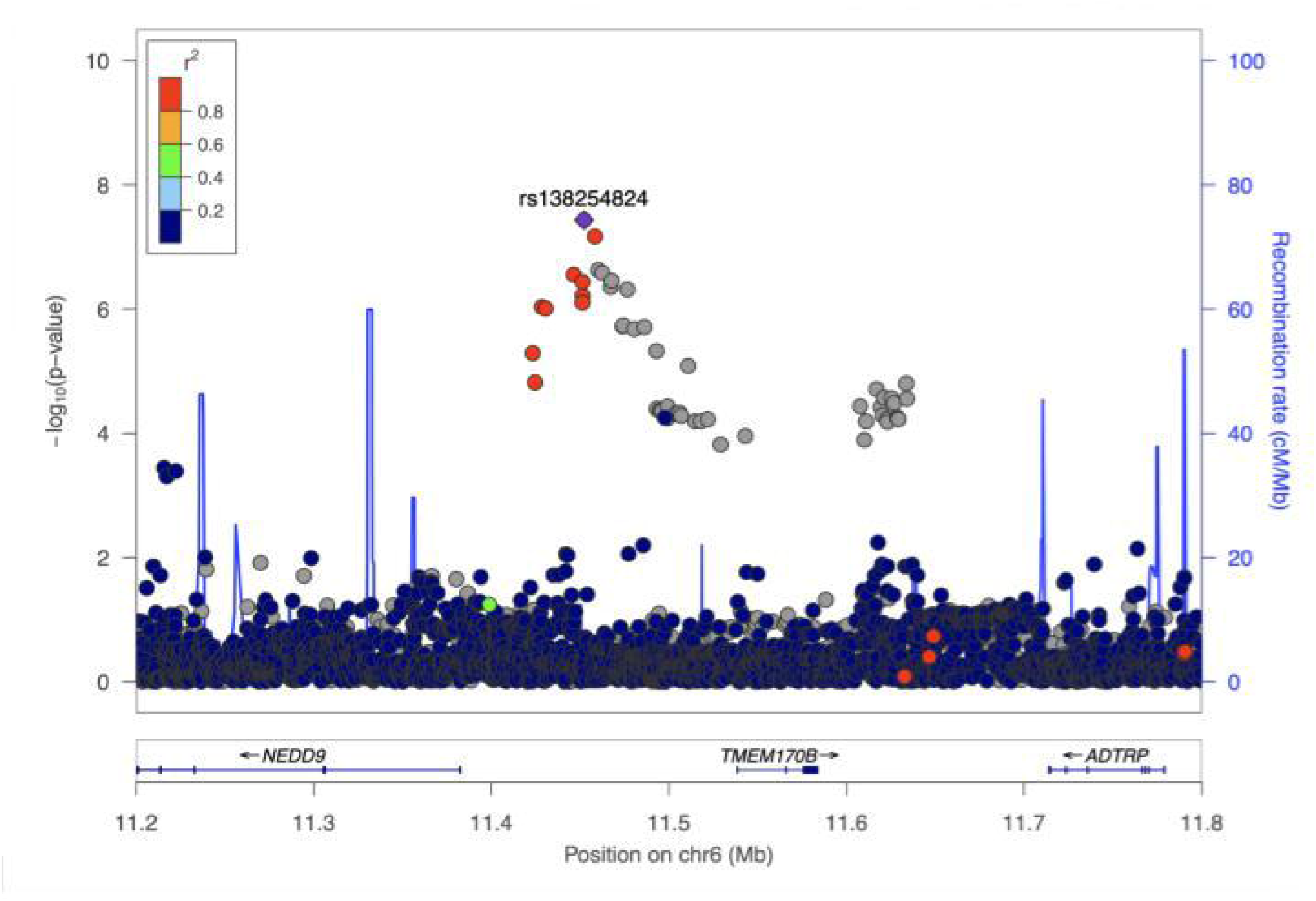
Regional-association plots for loci on chromosome 4 (A) and chromosome 6 (B). Tested SNPs are arranged by genomic position on chromosome 4 (A) or chromosome 6 (B) on the x-axis. The y-axis indicates −log10 p-values for association with lateral epicondylopathy. The color of dots representing flanking SNPs indicates their linkage disequilibrium (r2) with the lead SNP as indicated in the heat map color key.

### Functional analysis of the lateral epicondylopathy loci

None of the 144 SNPs on chromosome 4 are located in a coding region, so they cannot affect protein function directly. We asked whether they may affect the expression of nearby genes. First, data from Regulomedb indicates that rs13127477 is located in a region of open chromatin, which is a property associated with active gene expression. Second, data from the Genotype-Tissue Expression (GTEX) portal indicate that rs13127477 is associated with expression of three nearby genes (*IBSP, MEPE* and *SPP1)*. Specifically, the G allele of rs13127477 is associated with increased expression of *IBSP* (19%), *MEPE* (17%) and *SPP1* (29%)(Figure 4). GWAS data from this study indicate that the G allele of rs13127477 is also associated with increased risk for lateral epicondylopathy, implying that higher expression of these gene(s) increases the risk for lateral epicondylopathy. The effect of the G allele could be due to increased expression of all three genes acting synergistically or it could be due to just one or two of the genes, in which case the other genes may not have a functional role regarding lateral epicondylopathy.

**Fig 4.**
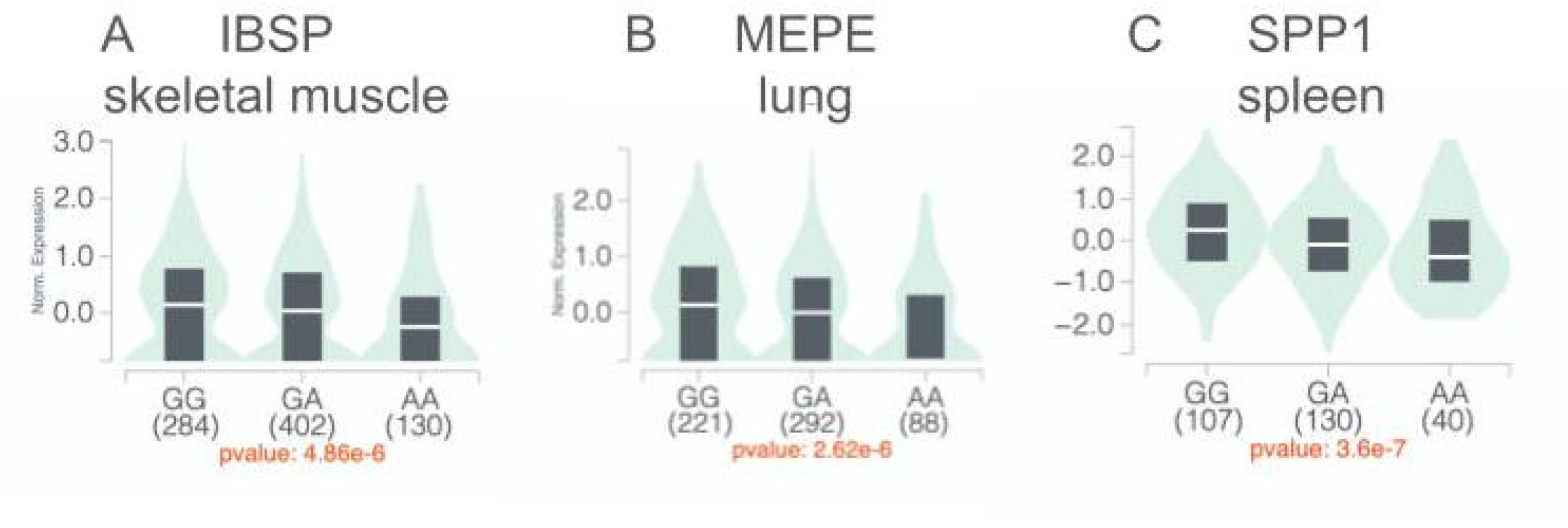
rs13127477 is associated with the expression of *IBSP, MEPE* and *SPP1*. Shown are violin plots for the effect of rs13127477 on gene expression. The gene and tissue that was assayed are shown on top. The expression level of the gene for each genotype is shown by the violin, where the width of the violin indicates the kernel density estimate of normalized expression. The black rectangle indicates the interquartile range of expression, with the median level shown by a horizontal bar. The x-axis shows the genotype that was assayed with the number of individuals with each genotype shown in parentheses. The y-axis shows the normalized expression size. The p-value that the SNP is an expression quantitative trait is shown at the bottom. (A) IBSP expression in skeletal muscle (B) MEPE expression in lung (C) SPP1 expression in spleen.

The SNP on chromosome 6 (rs138254824) lies between the genes *NEDD9* and *TMEM170B* (Fig 3B). The NEDD9 (Neural Precursor Cell Expressed, Developmentally Down-Regulated 9) scaffold protein is involved in transduction of extracellular matrix–integrin signals that guide osteoclast motility.^15^ TMEM170B (Transmembrane Protein 170B) is a transmembrane regulatory protein involved in Wnt/β-catenin signaling that is involved in bone formation pathways.^16^ rs138254824 is not located within a protein coding region nor is it known to be associated with changes in expression of these genes according to the GTEX portal. Thus, it is currently unclear how this polymorphism affects risk for lateral epicondylopathy.

To gain further biological insight into how the SNPs may affect risk for lateral epicondylopathy, we queried whether any of the linked SNPs reported herein were reported in previous GWA studies (Table 4). Two studies found that two SNPs from the chromosome 4 locus (rs5860110 and rs10011284) were associated with levels of osteopontin, encoded by the *SPP1* gene.^17,18^ Two other SNPs from the chromosome 4 locus (rs10013836 and rs2728127) were found to be associated with levels of urate and risk for gout.^19,20^ One study found that rs6833161 was associated with levels of oleoyl-arachidonoyl-glycerol levels in elite athletes.^21^ Finally, one study found that rs1381949 was associated with elevated blood protein levels.^22^ All of these SNPs are highly correlated with the lead SNP rs13127477, indicating that it is likely that it is also associated with these traits.

**Table 4:**
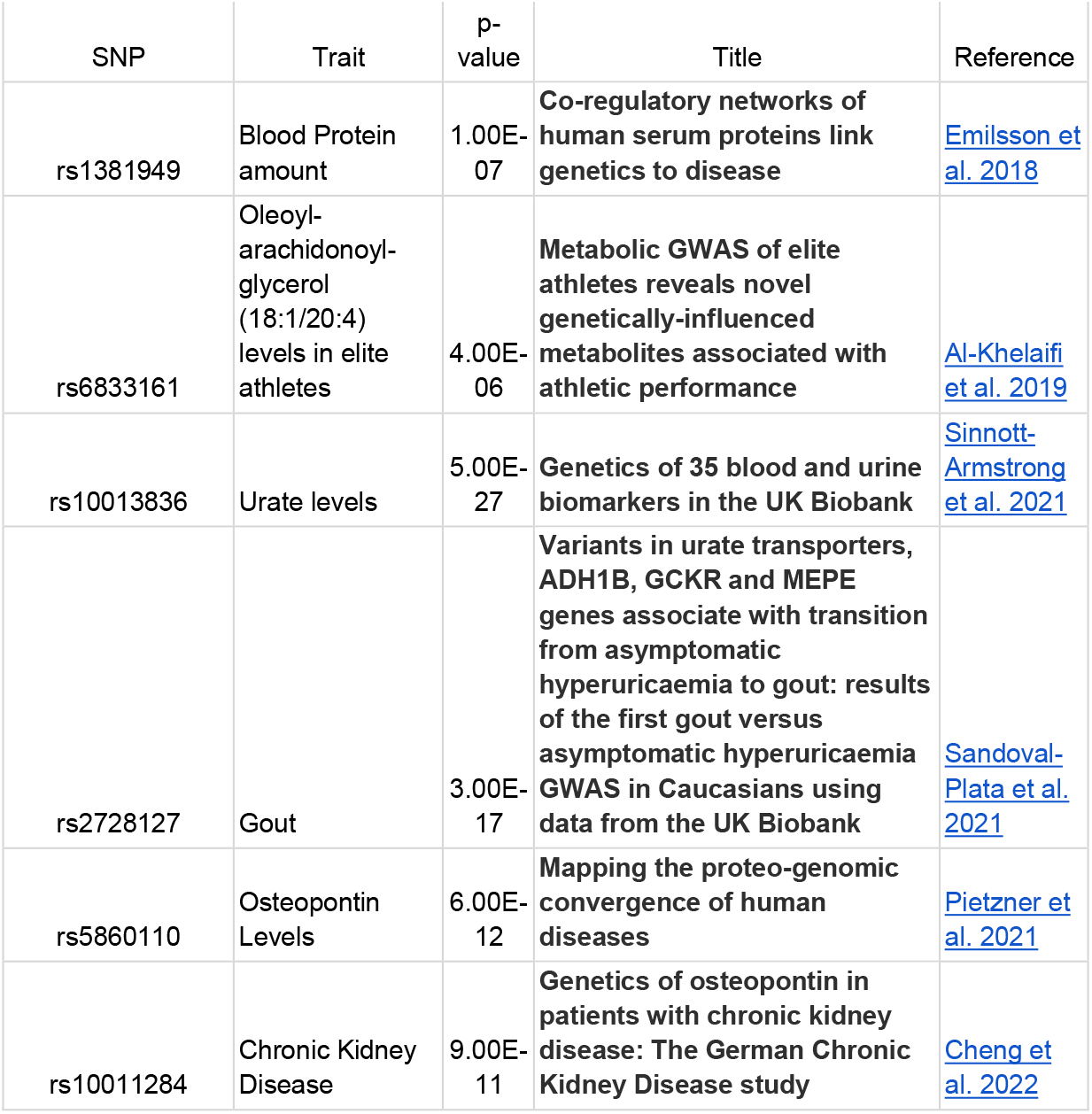
Pleiotropic effects of the SNPs for lateral epicondylopathy.

| SNP | Trait | p-value | Title | Reference |
| --- | --- | --- | --- | --- |
| rs1381949 | Blood Protein amount | 1.00E-07 | **Co-regulatory networks of human serum proteins link genetics to disease** | Emilsson et al. 2018 |
| rs6833161 | Oleoyl-arachidonoyl-glycerol (18:1/20:4) levels in elite athletes | 4.00E-06 | **Metabolic GWAS of elite athletes reveals novel genetically-influenced metabolites associated with athletic performance** | Al-Khelaifi et al. 2019 |
| rs10013836 | Urate levels | 5.00E-27 | **Genetics of 35 blood and urine biomarkers in the UK Biobank** | Sinnott-Armstrong et al. 2021 |
| rs2728127 | Gout | 3.00E-17 | **Variants in urate transporters, ADH1B, GCKR and MEPE genes associate with transition from asymptomatic hyperuricaemia to gout: results of the first gout versus asymptomatic hyperuricaemia GWAS in Caucasians using data from the UK Biobank** | Sandoval-Plata et al. 2021 |
| rs5860110 | Osteopontin Levels | 6.00E-12 | **Mapping the proteo-genomic convergence of human diseases** | Pietzner et al. 2021 |
| rs10011284 | Chronic Kidney Disease | 9.00E-11 | **Genetics of osteopontin in patients with chronic kidney disease: The German Chronic Kidney Disease study** | Cheng et al. 2022 |

### Failure to validate previous candidate gene studies

Previous studies have reported that polymorphisms in candidate collagen genes *col5A1* (rs13946, rs12722) and *col11A1* (rs3753841) are associated with lateral epicondylopathy.^4,5^ In our analysis, none of these SNPs showed an association with lateral epicondylopathy, even though the study size in the work presented here is much larger than in the previous candidate gene studies (20,390 vs 137 or 152, respectively). Specifically, rs13946, rs12722 and rs3753841 had p-values for lateral epicondylopathy of 0.99, 0.19 and 0.23 in our dataset. Thus, we were unable to replicate the previous candidate gene results for lateral epicondylopathy.

## Discussion

This study provides the first genome-wide analysis of lateral epicondylopathy, identifying two associated loci. The primary locus on chromosome 4 maps to three SIBLING genes—IBSP, MEPE, and SPP1—which regulate biomineralisation, entheseal structure, and extracellular matrix remodelling at the fibrocartilaginous enthesis, a key site in lateral epicondylopathy.^3,14^ These findings link genetic susceptibility to biological processes relevant to tendon-bone interface pathology and recovery.

The chromosome 4 locus encompasses three nearby genes: *IBSP, MEPE* and *SPP1. IBSP* mediates integrin-dependent cell adhesion, migration and matrix mineralization and influences osteoblast and osteoclast activity at the bone-tendon interface under repetitive loading.^23,24^ MEPE regulates bone turnover, mineralization and vascularization, processes relevant to tendon repair capacity and neovascular responses observed in tendinopathy.^25^ *SPP1* has roles in cell adhesion and chemotaxis, which play roles in fibroblast infiltration, vascular hyperplasia, and disorganised collagen deposition characteristic of angiofibroblastic tendinosis.^24,26^ Consistent with these functions, the risk allele was associated with increased expression of all SIBLING genes (Fig. 4), suggesting that excessive remodelling and pathological matrix regulation contribute to disease susceptibility.

The second locus on chromosome 6 lies near *NEDD9* and *TMEM170B. NEDD9* regulates FAK– SRC signalling, integrin activation, and focal adhesion dynamics, key pathways in cellular mechanosensing and tendon microtrauma repair, and has also been implicated in osteoclastogenesis relevant to bone remodelling at the tendon–bone interface.^27,28^ *TMEM170B* participates in Wnt/β-catenin signalling, a pathway critical for bone formation and maintenance that may influence entheseal adaptation to mechanical loading.^29,30^

### Pleiotropic effects of SIBLING genes

The chromosome 4 locus demonstrates pleiotropic associations, as the same SNPs have been found in previous studies for different traits (Table 4). Variants at this locus (rs5860110 and rs10011284) have been associated with circulating levels of osteopontin levels, consistent with the expression findings in Figure 4.^17,18^ Second, rs6833161 is associated with levels of oleoyl-arachidonoyl-glycerol (18:1/20:4) in elite athletes.^21^ Oleoyl-arachidonoyl-glycerol is a type of diacylglycerol, which is known to be reduced in high endurance athletes.^31^ Third, rs2728127 and rs10013836 have been associated with serum urate levels and gout risk, respectively.^19,20^ Tendons and entheses are increasingly recognised as sites of monosodium urate deposition, which may present as non-specific regional pain rather than classic inflammatory arthritis.^32,33^

### Previous candidate gene studies

Previous studies have tested two collagen genes (*COL5A1* and *COL11A1*) as candidates for association with lateral epicondylopathy.^4,5^ Our GWAS contains many more cases of lateral epicondylopathy (20,390) than were available in the previous studies (152 and 137, respectively). We performed follow-up experiments using the GWAS results in an attempt to validate the results, and were not able to replicate any of the previous reports. Power calculations indicate an 80% chance of success if the genotype relative risk of these candidate genes were at least 1.03. One explanation for the lack of validation is that the previous studies looked at cases of lateral epicondylopathy in athletes, whereas our study looked at individuals from the general population. Nevertheless, evidence from many other studies suggests that candidate gene associations need to be independently replicated; otherwise, their credibility is low.^34^

### Personalized Treatment

The identification of genetic loci associated with lateral epicondylopathy may inform future risk stratification and personalized treatment strategies. Variants affecting SIBLING gene regulation implicate pathways involved in entheseal extracellular matrix remodelling, highlighting biological mechanisms relevant to tendon–bone interface adaptation and repair.

Platelet rich plasma (PRP) therapy for lateral epicondylopathy demonstrate significant heterogeneity in treatment outcomes, with variability attributed to differences in PRP formulation, platelet concentration, and patient specific biologic factors.^4,35^ Emerging research suggests that polymorphisms of VEGF, TGFB1, PDGFB are associated with differential PRP response with specific genotypes demonstrating better outcomes over two-year follow up.^36,37^ Notably, these factors directly regulate SIBLING protein expression which may suggest a mechanistic intersection where genetic variation could influence clinical outcomes.

### Limitations and Future studies

Several limitations should be considered when interpreting these findings. Case definition relied on diagnostic codes taken from electronic health records (EHR) rather than standardized clinical assessments. Using standardized clinical assessments may reduce misclassifications related to variable inclusion criteria coding practices, variability between providers, or under-recognized cases. The analysis was restricted to individuals of European ancestry, limiting generalisability to other populations. Additionally, as this represents the first genome-wide screen for lateral epicondylopathy, independent replication in external cohorts is required.

Future studies should evaluate these loci in athletic populations and investigate whether genotyping for variants affecting *SPP1, IBSP, MEPE* can help identify individuals more likely to benefit from biologically targeted interventions such as PRP. Such work may support precision prehabilitation and treatment strategies, particularly in athletic populations where injury prevention, durability of recovery, and recurrence reduction are critical.

## Data Availability

All data produced in the present study are available upon reasonable request to the authors

## Footnotes

### Ethical approval

This study analyzed stored data from UK Biobank subjects who consented to genomic testing and use of their genomic data, as well as health data from the UK Biobank electronic health records. The health and genotype data for the subjects were deidentified.

### Patient involvement

Patients were not involved in this research, except as anonymized subjects in the two cohorts.

### Contributorship

KB, SKK, and WD contributed equally to the conception and design of the study, data analysis and interpretation, and drafting and critical revision of the manuscript. All authors approved the final manuscript. WD is the guarantor.

## Acknowledgements

The authors thank Chris Chang for Plink, and the UK Biobank and GTEX for access to data

## Funding, grant and award information

This research received no specific grant from any funding agency in the public, commercial, or not-for-profit sectors.

## Equity, Diversity, and Inclusion

This study analysed UK Biobank data and was restricted to participants of European ancestry to reduce population stratification, limiting generalisability to other groups and underscoring the need for future studies in more diverse populations.

## References

1. Field LD, Savoie FH. Common elbow injuries in sport. Sports Med. 1998 Sept;26(3):193–205.

2. Physical therapy for people with lateral elbow tendinopathy: Using the evidence to guide musculoskeletal rehabilitation clinical practice. J Orthop Sports Phys Ther. 2023 Jan;53(1):5–6.

3. Wolf JM. Lateral epicondylitis. N Engl J Med. 2023 June 22;388(25):2371–7.

4. Alakhdar Mohmara Y, Cook J, Benítez-Martínez JC, McPeek ER, Aguilar AA, Olivas ES, et al. Influence of genetic factors in elbow tendon pathology: a case-control study. Sci Rep. 2020 Apr 16;10(1):6503.

5. Altinisik J, Meric G, Erduran M, Ates O, Ulusal AE, Akseki D. The BstUI and DpnII variants of the COL5A1 gene are associated with tennis elbow. Am J Sports Med. 2015 July;43(7):1784–9.

6. Bycroft C, Freeman C, Petkova D, Band G, Elliott LT, Sharp K, et al. The UK Biobank resource with deep phenotyping and genomic data. Nature. 2018 Oct;562(7726):203–9.

7. Howie B, Marchini J, Stephens M. Genotype imputation with thousands of genomes. G3 Bethesda. 2011 Nov;1(6):457–70.

8. Chang CC, Chow CC, Tellier LC, Vattikuti S, Purcell SM, Lee JJ. Second-generation PLINK: rising to the challenge of larger and richer datasets. Gigascience. 2015 Feb 25;4(1):7.

9. Pruim RJ, Welch RP, Sanna S, Teslovich TM, Chines PS, Gliedt TP, et al. LocusZoom: regional visualization of genome-wide association scan results. Bioinformatics. 2010;26(18):2336–7.

10. Boyle AP, Hong EL, Hariharan M, Cheng Y, Schaub MA, Kasowski M, et al. Annotation of functional variation in personal genomes using RegulomeDB. Genome Res. 2012;22(9):1790–7.

11. Ganss B, Kim RH, Sodek J. Bone sialoprotein. Crit Rev Oral Biol Med. 1999;10(1):79–98.

12. Rowe PSN, Kumagai Y, Gutierrez G, Garrett IR, Blacher R, Rosen D, et al. MEPE has the properties of an osteoblastic phosphatonin and minhibin. Bone. 2004 Feb;34(2):303–19.

13. Si J, Wang C, Zhang D, Wang B, Zhou Y. Osteopontin in bone metabolism and bone diseases. Med Sci Monit. 2020 Jan 30;26:e919159.

14. Staines KA, MacRae VE, Farquharson C. The importance of the SIBLING family of proteins on skeletal mineralisation and bone remodelling. J Endocrinol. 2012 Sept;214(3):241–55.

15. Nikonova AS, Gaponova AV, Kudinov AE, Golemis EA. CAS proteins in health and disease: an update: CAS Proteins: Recent Developments. IUBMB Life. 2014 June;66(6):387–95.

16. Herrera-Quiterio GA, Encarnación-Guevara S. The transmembrane proteins (TMEM) and their role in cell proliferation, migration, invasion, and epithelial-mesenchymal transition in cancer. Front Oncol. 2023 Oct 23;13:1244740.

17. Cheng Y, Li Y, Scherer N, Grundner-Culemann F, Lehtimäki T, Mishra BH, et al. Genetics of osteopontin in patients with chronic kidney disease: The German Chronic Kidney Disease study. PLoS Genet. 2022 Apr;18(4):e1010139.

18. Pietzner M, Wheeler E, Carrasco-Zanini J, Cortes A, Koprulu M, Wörheide MA, et al. Mapping the proteo-genomic convergence of human diseases. Science. 2021 Nov 12;374(6569):eabj1541.

19. Sinnott-Armstrong N, Tanigawa Y, Amar D, Mars N, Benner C, Aguirre M, et al. Genetics of 35 blood and urine biomarkers in the UK Biobank. Nat Genet. 2021 Feb;53(2):185–94.

20. Sandoval-Plata G, Morgan K, Abhishek A. Variants in urate transporters, ADH1B, GCKR and MEPE genes associate with transition from asymptomatic hyperuricaemia to gout: results of the first gout versus asymptomatic hyperuricaemia GWAS in Caucasians using data from the UK Biobank. Ann Rheum Dis. 2021 Sept;80(9):1220–6.

21. Al-Khelaifi F, Diboun I, Donati F, Botrè F, Abraham D, Hingorani A, et al. Metabolic GWAS of elite athletes reveals novel genetically-influenced metabolites associated with athletic performance. Sci Rep. 2019 Dec 27;9(1):19889.

22. Emilsson V, Ilkov M, Lamb JR, Finkel N, Gudmundsson EF, Pitts R, et al. Co-regulatory networks of human serum proteins link genetics to disease. Science. 2018 Aug 24;361(6404):769–73.

23. Bouleftour W, Juignet L, Bouet G, Granito RN, Vanden-Bossche A, Laroche N, et al. The role of the SIBLING, Bone Sialoprotein in skeletal biology - Contribution of mouse experimental genetics. Matrix Biol. 2016 May;52–54:60–77.

24. Marinovich R, Soenjaya Y, Wallace GQ, Zuskov A, Dunkman A, Foster BL, et al. The role of bone sialoprotein in the tendon-bone insertion. Matrix Biol. 2016 May;52–54:325–38.

25. Rowe PS. A unified model for bone-renal mineral and energy metabolism. Curr Opin Pharmacol. 2015 June;22:64–71.

26. Whyte MP, Amalnath SD, McAlister WH, McKee MD, Veis DJ, Huskey M, et al. Hypophosphatemic osteosclerosis, hyperostosis, and enthesopathy associated with novel homozygous mutations of DMP1 encoding dentin matrix protein 1 and SPP1 encoding osteopontin: The first digenic SIBLING protein osteopathy? Bone. 2020 Mar;132(115190):115190.

27. Shagisultanova E, Gaponova AV, Gabbasov R, Nicolas E, Golemis EA. Preclinical and clinical studies of the NEDD9 scaffold protein in cancer and other diseases. Gene. 2015 Aug 1;567(1):1–11.

28. Zhong J, Baquiran JB, Bonakdar N, Lees J, Ching YW, Pugacheva E, et al. NEDD9 stabilizes focal adhesions, increases binding to the extra-cellular matrix and differentially effects 2D versus 3D cell migration. PLoS One. 2012 Apr 11;7(4):e35058.

29. Astudillo P, Larraín J. Wnt signaling and cell-matrix adhesion. Curr Mol Med. 2014 Feb;14(2):209–20.

30. Duan P, Bonewald LF. The role of the wnt/β-catenin signaling pathway in formation and maintenance of bone and teeth. Int J Biochem Cell Biol. 2016 Aug;77(Pt A):23–9.

31. Al-Khelaifi F, Diboun I, Donati F, Botrè F, Alsayrafi M, Georgakopoulos C, et al. A pilot study comparing the metabolic profiles of elite-level athletes from different sporting disciplines. Sports Med Open. 2018 Jan 5;4(1):2.

32. McGonagle D, Lories RJU, Tan AL, Benjamin M. The concept of a “synovio-entheseal complex” and its implications for understanding joint inflammation and damage in psoriatic arthritis and beyond. Arthritis Rheum. 2007 Aug;56(8):2482–91.

33. Dalbeth N, Horne A, Gamble GD, Ames R, Mason B, McQueen FM, et al. The effect of calcium supplementation on serum urate: analysis of a randomized controlled trial. Rheumatol Oxf Engl. 2009 Feb;48(2):195–7.

34. Siontis KC, Patsopoulos NA, Ioannidis JP. Replication of past candidate loci for common diseases and phenotypes in 100 genome-wide association studies. Eur J Hum Genet. 2010;18(7):832–7.

35. Lu J, Li H, Zhang Z, Xu R, Wang J, Jin H. Platelet-rich plasma in the pathologic processes of tendinopathy: a review of basic science studies. Front Bioeng Biotechnol. 2023;11:1187974.

36. Niemiec P, Jarosz A, Balcerzyk-Matić A, Iwanicka J, Nowak T, Iwanicki T, et al. Genetic Variability in VEGFA Gene Influences the Effectiveness of Tennis Elbow Therapy with PRP: A Two-Year Prospective Cohort Study. Int J Mol Sci. 2023 Dec 9;24(24):17292.

37. Jarosz A, Wrona J, Balcerzyk-Matić A, Szyluk K, Nowak T, Iwanicki T, et al. Association of the TGFB1 Gene Polymorphisms with Pain Symptoms and the Effectiveness of Platelet-Rich Plasma in the Treatment of Lateral Elbow Tendinopathy: A Prospective Cohort Study. Int J Mol Sci. 2025 Mar 8;26(6):2431.

